# Systematic review and network meta-analysis with individual participant data on Cord Management at Preterm Birth (iCOMP): study protocol

**DOI:** 10.1101/19007708

**Authors:** Anna Lene Seidler, Lelia Duley, Anup Katheria, Catalina De Paco Matallana, Eugene Dempsey, Heike Rabe, John Kattwinkel, Judith Mercer, Justin Josephsen, Karen Fairchild, Ola Andersson, Shigeharu Hosono, Venkataseshan Sundaram, Vikram Datta, Walid El-Naggar, William Tarnow-Mordi, Thomas P.A. Debray, Stuart Hooper, Martin Kluckow, Graeme Polglase, Peter Davis, Alan Montgomery, Kylie E Hunter, Angie Barba, John Simes, Lisa Askie, on behalf of the iCOMP collaboration

## Abstract

**Introduction:** Timing of cord clamping and other cord management strategies may improve outcomes at preterm birth. However, it is unclear whether benefits apply to all preterm subgroups such as those who usually receive immediate neonatal care. Previous and current trials compare various policies, including immediate cord clamping, time- or physiology-based deferred cord clamping, and cord milking. Individual participant data (IPD) enables exploration of different strategies within subgroups. Network meta-analysis (NMA) enables comparison and ranking of all available interventions using a combination of direct and indirect comparisons.

**Objectives:** 1) To evaluate the effectiveness of cord management strategies for preterm infants on neonatal mortality and morbidity overall and for different participant characteristics using IPD meta-analysis; and 2) to evaluate and rank the effect of different cord management strategies for preterm births on mortality and other key outcomes using NMA.

**Methods and analysis:** We will conduct a systematic search of Medline, Embase, clinical trial registries, and other sources for all planned, ongoing and completed randomised controlled trials comparing alternative cord management strategies at preterm birth (before 37 weeks’ gestation). IPD will be sought for all trials. First, deferred clamping and cord milking will be compared with immediate clamping in pairwise IPD meta-analyses. The primary outcome will be death prior to hospital discharge. Effect differences will be explored for pre-specified subgroups of participants. Second, all identified cord management strategies will be compared and ranked in an IPD NMA for the primary outcome and the key secondary outcomes intraventricular haemorrhage (any grade) and infant blood transfusions (any). Treatment effect differences by participant characteristics will be identified. Inconsistency and heterogeneity will be explored.

**Ethics and dissemination:** Approved by University of Sydney Human Research Ethics Committee (2018/886). Results will be relevant to clinicians, guideline-developers and policy-makers, and will be disseminated via publications, presentations, and media releases.

**Registration:** Australian New Zealand Clinical Trials Registry: ACTRN12619001305112.

**STRENGTH AND LIMITATIONS OF THIS STUDY:**

- This will be the most comprehensive review to date of interventions for umbilical cord management in preterm infants and the findings will be highly relevant to clinicians and guideline developers
- The use of individual participant data will allow assessment of the best treatment option for key subgroups of participants
- Network meta-analysis will enable the comparison and ranking of all available treatment options using direct and indirect evidence
- For some of the trials it will not be possible to obtain individual participant data, so published aggregate results will be used instead
- Risk of bias in the primary trials will be assessed using Cochrane criteria, and certainty of evidence for the meta-analyses will be appraised using the GRADE approach for the pairwise comparisons, and the CINeMA approach for the network meta-analysis

## INTRODUCTION

Currently over 15 million babies are born preterm annually and this number is rising.(1-3) Of these, 1.1 million die, and the morbidity and healthcare costs amongst survivors and their families are high, with preterm survivors having an increased risk of cognitive, developmental and behavioural difficulties, and chronic ill health.(4-9) Hence, even modest improvements in outcomes of preterm birth would substantially benefit the children, their families, and also health services. In uncompromised babies, deferring cord clamping has been shown to be beneficial and is now used in routine practice.(10) However, it is unclear whether these benefits apply to preterm babies who usually receive immediate neonatal care, and whether any benefits outweigh potential harms. In addition, there are multiple competing cord management strategies, such as clamping the cord at different times or milking the cord, and considerations of the infant’s respiratory status, and it is currently unknown which strategy yields the best balance of benefits and harms.

### Current approaches to cord clamping

One potential mechanism of deferring umbilical cord clamping is a net transfer of blood from the placenta to the baby known as “placental transfusion”. If the cord is not clamped at birth immediately, blood flow between the placenta and the baby may continue for up to five minutes in term infants.(11-13) For preterm births, blood flow may continue for longer,(14) since a greater proportion of feto-placental circulating blood volume is still in the placenta.(15) This has led to time-based approaches to deferring cord clamping that have been shown to increase peak haematocrit and reduce the need for blood transfusions.(16) Yet, recent findings suggest that placental transfusion does not always occur - blood flow may continue without any net transfer, and sometimes net transfer may be to the placenta.(17) Initial neonatal care and stabilisation traditionally takes place on a resuscitation platform at the side of the room or in an adjacent room. Deferred cord clamping is thus often associated with a delay in neonatal care and this has led to concerns including delayed resuscitation and hypothermia (18) particularly for very preterm infants and infants assessed as requiring resuscitation. An alternate emerging strategy is to provide immediate neonatal care with the cord intact beside the woman using a mobile resuscitation trolley or on the mother’s leg. (19-24)

Another potential mechanism of deferred clamping is allowing time for the infant to establish spontaneous breathing whilst still placentally supported. Immediate cord clamping before the infant has established breathing may be harmful since it can lead to large fluctuations in blood pressure, a period of hypoxia, and restricted cardiac function.(25) Animal and pilot human studies suggest that breathing and lung aeration before cord clamping can improve cardiovascular stability and oxygenation, and reduce intraventricular haemorrhage and infant mortality.(26-29) They also suggest that initial respiratory support before clamping the cord can improve cerebral oxygenation and blood pressure, and reduce cerebrovascular impairment compared with immediate cord clamping.(30, 31) This evidence has led to the rise of “physiological cord clamping” which defers clamping until after the onset of breathing. Yet, onset of breathing is not always easy to determine without the assistance of video or extra equipment, whilst timing to cord clamping can be easily measured. In an earlier study,(32) time of onset of breathing in preterm infants receiving gentle stimulation was related to time after birth – within a minute over 90% of preterm infants had begun spontaneous breathing.

Cord milking or stripping (pinching the umbilical cord close to the mother and moving the fingers towards the infant) may be a way to increase preterm blood volume without deferring clamping.(33) Yet, a preterm lamb model demonstrated that during cord milking there was a transient increase to carotid blood flow and pressure.(34) A recent trial comparing deferred cord clamping with cord milking was stopped early in the subgroup of extremely preterm infants (23-27 weeks), as the incidence of severe intraventricular haemorrhage was higher in the cord milking group.(35) Hence, the effect of cord milking in different populations needs further elucidation.

### Current guidelines for cord management at birth and previous reviews of aggregate data

Current uncertainties in optimal cord management strategies are reflected in varying guidelines. The World Health Organization (WHO) recommends late cord clamping (36) unless resuscitation is required, the National Institute for Health and Care Excellence (NICE) recommends waiting for 30 seconds to 3 minutes if mother and baby are stable, (37) and the International Liaison Committee on Resuscitation Council (ILCOR) recommends a delay in cord clamping of at least 1 minute. If the baby is assessed as requiring resuscitation (which is the case in many preterm infants), (38) WHO recommends immediate clamping, (39) NICE recommends considering cord milking before clamping, and ILCOR concludes that there is insufficient evidence to make any recommendations. (38)

A 2012 Cochrane review of timing of cord clamping for preterm births (40) included 15 trials, with 738 infants, of which one trial (with 40 infants) compared cord milking with immediate cord clamping.(41) There was heterogeneity in the timing of cord clamping and gestational age at recruitment, and data were insufficient for reliable conclusions about any of the primary outcomes of the review. A systematic review and meta-analysis published in 2018 (including 18 trials with 2834 infants) compared the effect of deferred (≥30 seconds) versus early (<30 seconds) clamping in preterm infants, and found a reduction in the primary outcome of hospital mortality by 32% (Risk Ratio = 0.68, 95% Confidence Interval = 0.52-0.90). (16) There was heterogeneity in the definition of ‘early cord clamping’ ranging from less than 5 to 25 seconds, and “late cord clamping”, ranging from 30 to 180 seconds. Recruitment age varied from 22 weeks to 36 weeks gestational age. Most analyses of infant and maternal morbidity were substantially underpowered. (16) The review concluded that while there is high quality evidence that deferred cord clamping improves outcomes, individual participant data analyses are urgently needed to further understand the benefits and potential harms of different cord management strategies, and to understand whether differential treatment options are advantageous for key subgroups of infants. (16)

This ongoing uncertainty about the optimal cord management strategy, and differential cord management strategies for key subgroups of infants (e.g. for those for which resuscitation and/or stabilisation is deemed necessary, or extremely preterm infants) has led to 113 planned, ongoing or published trials (in more than 15,000 preterm babies) that are comparing a range of cord management strategies. *Individual participant data (IPD) meta-analysis* is the gold standard for combining such trial data. IPD provides larger statistical power for estimation of treatment effects of rarer secondary endpoints and enables reliable subgroup analyses to examine hypotheses about differences in treatment effect, exploring interactions between treatment- and participant-level characteristics. (42) A *network meta-analysis (NMA)* facilitates data synthesis when there are a range of interventions available and permits indirect comparisons across all interventions by inferring the relative effectiveness of two competing treatments through a common comparator.(43, 44) NMA produces relative effect estimates for each intervention compared with every other intervention in the network. These effect sizes can be used to obtain rankings of the effectiveness of the interventions.(45) Using IPD in a NMA (as opposed to aggregate data) can improve precision, increase information, and reduce bias. (46)

### Objectives

The aims of this study are:

1. to evaluate the effectiveness of cord management strategies for preterm infants on neonatal mortality and morbidity, and to evaluate patient-level modifiers of treatment effect.
2. to evaluate, compare and rank the effectiveness of different cord management strategies for preterm infants on mortality and the key secondary outcomes intraventricular haemorrhage (any grade) and infant blood transfusions (any).

## METHODS AND ANALYSIS

We will conduct a systematic review of randomised trials with individual participant data using pairwise and network meta-analysis, and a nested prospective meta-analysis. The lead investigator for all potentially eligible studies will be contacted and invited to collaborate and join the **<u>i</u>**<u>ndividual participant data</u> **<u>Co</u>**<u>rd</u> **<u>M</u>**<u>anagement at</u> **<u>P</u>**<u>reterm birth (iCOMP) Collaboration</u>. Eligible trials identified up to February 2019 are listed in Supplementary File 1. The Collaboration will undertake this project according to the methods recommended by the Cochrane Individual Participant Data, Multiple Interventions, and Prospective Meta-Analysis Methods Groups.(42, 47, 48) This protocol is registered on the Australian New Zealand Clinical Trials Registry (ANZCTR) and is undergoing editorial review for registration on the International Prospective Register of Systematic Reviews (PROSPERO). Reporting guidelines for NMA protocols by Chaimani et al (49) and PRISMA extension for IPDs and protocols (50, 51) have been followed for reporting (checklist provided in Supplementary File 2).

### Eligibility criteria

#### Types of studies

Studies will be included if they are randomised trials. Cluster-randomised and quasi-random studies will be excluded. Studies must compare at least two of the interventions of interest (defined below).

#### Trial participants

Participants will be women giving birth preterm (before 37 completed weeks’ gestation) and/or their babies. Individually randomised studies will be eligible for inclusion if the unit of randomisation was either the woman, or the baby. Women and babies will be included regardless of whether mode of delivery was vaginal or caesarean, and whether the birth was singleton or multiple. Babies will be included regardless of whether or not they received immediate resuscitation at birth.

#### Types of interventions and comparators in pairwise meta-analysis

For the pairwise meta-analysis we will include all trials that compare an intervention to enhance umbilical blood flow or allow more time for physiological transition to the comparator immediate cord clamping. This includes interventions assessing cord management strategies for timing of cord clamping, and other strategies such as cord milking. Trials will be included regardless of whether initial neonatal care is provided with the umbilical cord intact, or not. Different strategies (i.e. deferred cord clamping and cord milking) will be analysed in separate subgroups to assess comparability between the groups by assessing subgroup effects and heterogeneity. They will then be collapsed into one “cord management intervention” group if they are deemed comparable based on the previous subgroup assessments. If they are deemed non-comparable they will be analysed and interpreted separately.

#### Types of interventions and comparators in network meta-analysis

For the network meta-analysis we will include, as interventions of interest, strategies for timing of cord clamping, and other cord management strategies to influence umbilical flow and placental transfusion.

Thus, interventions of interest include:

- Immediate cord clamping without milking (≤15 seconds or trialist defined)
- Short deferral of cord clamping (>15 to <45 seconds) without milking
- Medium deferral of cord clamping (≥45 to <90 seconds) without milking
- Long deferral of cord clamping (≥ 90 seconds) without milking
- Cord milking or stripping before immediate cord clamping (intact cord milking)
- Cord milking or stripping before deferred cord clamping (intact cord milking)
- Cord milking or stripping after immediate cord clamping (cut cord milking)
- Cord milking or stripping after deferred cord clamping (cut cord milking)
- Physiological clamping (clamping after aeration of lungs)

If we identify other interventions not listed above we will include them if they are addressing cord management or related strategies to influence umbilical flow and placental transfusion. Again, trials will be included regardless of whether initial neonatal care is provided with the umbilical cord intact, or not. Studies evaluating collection and storage of residual placental blood that is then used for transfusion after birth will be excluded. All possible comparisons between eligible interventions are displayed in Figure 1. For interpretation purposes, immediate cord clamping will act as the basis comparison/ parameter.

**Figure 1.**
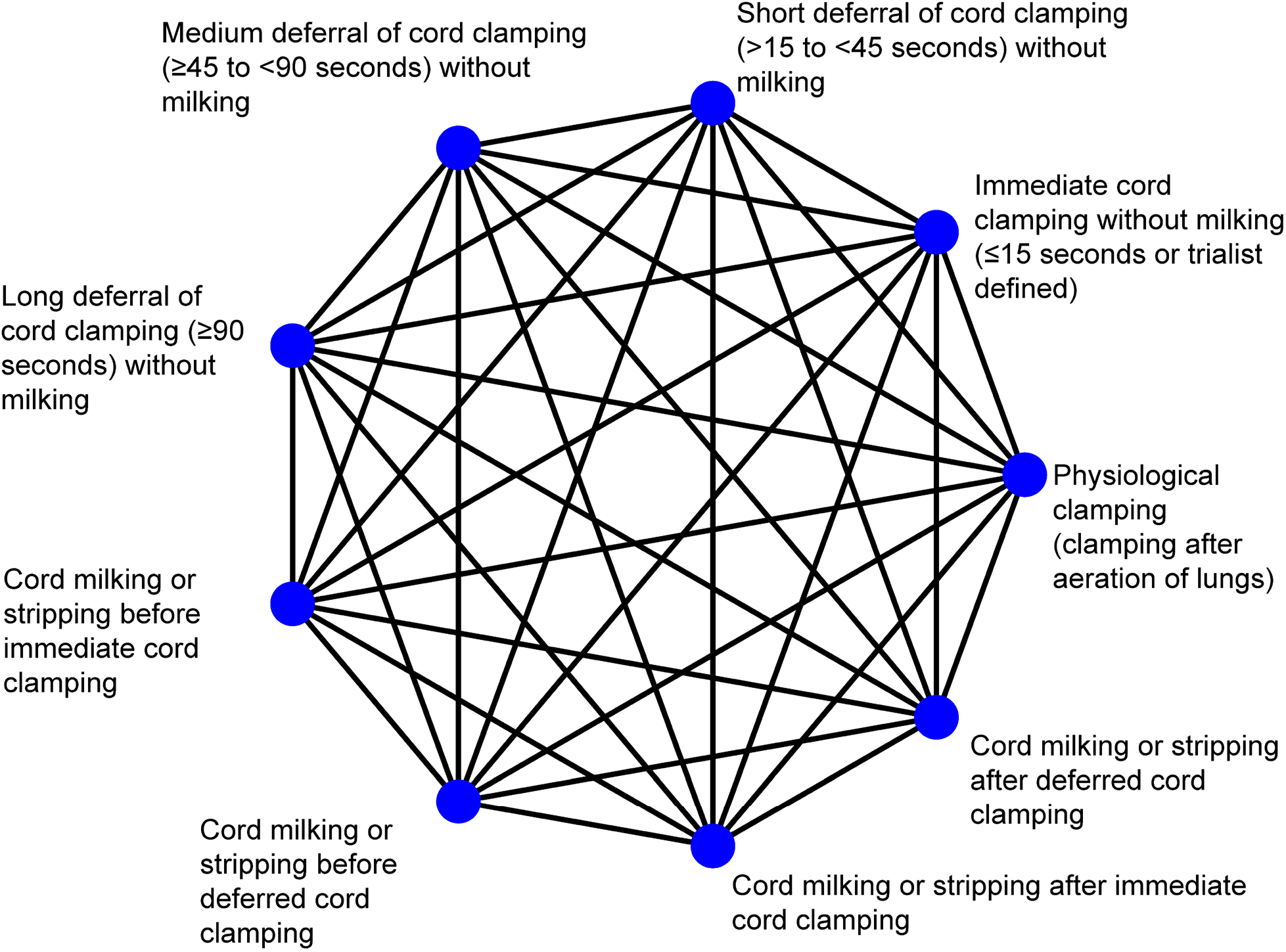
Network of possible comparisons between cord management interventions.

Nodes that specify different timings of cord clamping were defined according to what timing is classified as immediate clamping, short deferral, medium deferral or long deferral according to the literature to date (as shown in Supplementary File 1), and after discussion with clinicians. Different timings are commonly compared in head-to-head comparisons, hence, their classification as different intervention nodes. Similarly, nodes that specify cord milking were classified after a review of current milking techniques described in the literature and after discussion with clinicians. If insufficient data are available, categories will be collapsed where possible. For instance, milking before and after immediate cord clamping could be collapsed into one single immediate cord milking category, or medium and long delay could be collapsed into a medium to long delay category. We consider the interventions of interest to be jointly randomisable (i.e. each participant could, in principle, be randomised to any one of the interventions of interest).

#### Types of outcome measures

Trials must report at least one of the clinical outcomes included in this review, as specified in the “measures” section below, to be included.

#### Eligibility for nested prospective meta-analysis

Studies are only included in the nested prospective meta-analysis if the investigator/s were blind to outcome data by intervention group at the time the main components of the protocol (i.e. objectives, aims and hypotheses, eligibility criteria, subgroup and sensitivity analyses and main outcomes) were initially agreed in January 2015.

### Information sources and search strategy

The search strategy to identify potentially eligible studies will include a search of the Cochrane Collaboration Pregnancy and Childbirth Review Group’s Trial Register. This register contains trials identified from: monthly searches of the Cochrane Central Register of Controlled Trials (CENTRAL) and CINAHL (EBSCO); weekly searches of Medline (Ovid) and Embase (Ovid); hand searches of specialty journals and major conferences proceedings; and current awareness alerts from further journals and BioMed Central. Further details can be found elsewhere.(52) We will identify ongoing trials that may be eligible by searching for published protocols in Medline and Embase, searching online registries of clinical trials, and personal contacts (for example, by asking collaborators to notify any unregistered studies they are aware of). The Chief Investigators of eligible trials will be invited to join the iCOMP Collaboration. They will also be asked if they know of any further planned, ongoing or completed studies. The search strategy is outlined in more detail in Supplementary File 3.

#### Selection of studies for inclusion in the review

Two members of the iCOMP Secretariat (see project management section below) will independently assess all the potentially eligible studies identified for inclusion. Disagreements will be resolved by discussion or, if required, by consulting a third member of the iCOMP Secretariat. Studies that are not willing or able to provide IPD will be synthesised where possible using aggregate data.

### Data collection, management, and confidentiality

#### Data receipt

De-identified, individual participant level data will be provided by each participating trial. These data will be backed-up and stored in a centralised secure database.

#### Data processing

##### Data checking

For each trial, range and internal consistency of all variables will be checked. Intervention details and missing data will be checked against any protocols, published reports, and data collection sheets. Integrity of the randomisation process will be assessed by examining the chronological randomisation sequence and the balance of participant characteristics across intervention groups. Any inconsistencies or missing data will be discussed with the trialists and resolved by consensus. Each included study will be analysed separately and the results sent to the trial investigators for verification prior to inclusion in the iCOMP database. All trial-specific outcomes generated from the IPD will be cross-checked against published information via a series of crosstabs.

##### Data re-coding

Outcome data may have been collected in different formats across trials. Therefore, the de-identified data from each of the trials will be extracted and re-formatted into a commonly coded dataset.

##### Data transformation and collating

Once the data from each of the trials are finalised, they will be combined into a single dataset, but a trial identifier code for each participant will be retained. New variables will be generated from the combined dataset as required to address the hypotheses to be tested.

### Risk of bias assessment and certainty of evidence appraisal

Eligible studies will be assessed for risk of bias using the criteria described in the Cochrane Handbook(53): random sequence generation; allocation concealment; blinding of participants and personnel; blinding of outcome assessment; incomplete outcome data; selective reporting; and other bias. Certainty of evidence will be appraised using the GRADE approach (54) for the pairwise comparisons, and the rating approach suggested by Salanti and colleagues that is implemented in the CINeMA application for the network meta-analysis.(55)

### Outcomes measures for pairwise meta-analysis

All outcome measures for the pairwise meta-analysis are listed in Table 1. The primary outcome will be death of the baby prior to hospital discharge. As outcomes for babies born very preterm (before 32 weeks’ gestation) are different to those born moderately preterm (32 to 37 weeks), separate analyses will be conducted for these two groups of infants for the secondary outcomes. Where possible, definitions will be standardised, otherwise outcomes will be used as defined in individual trials. Secondary outcomes will include measures of neonatal and maternal morbidity, and health service use.

**Table 1.**
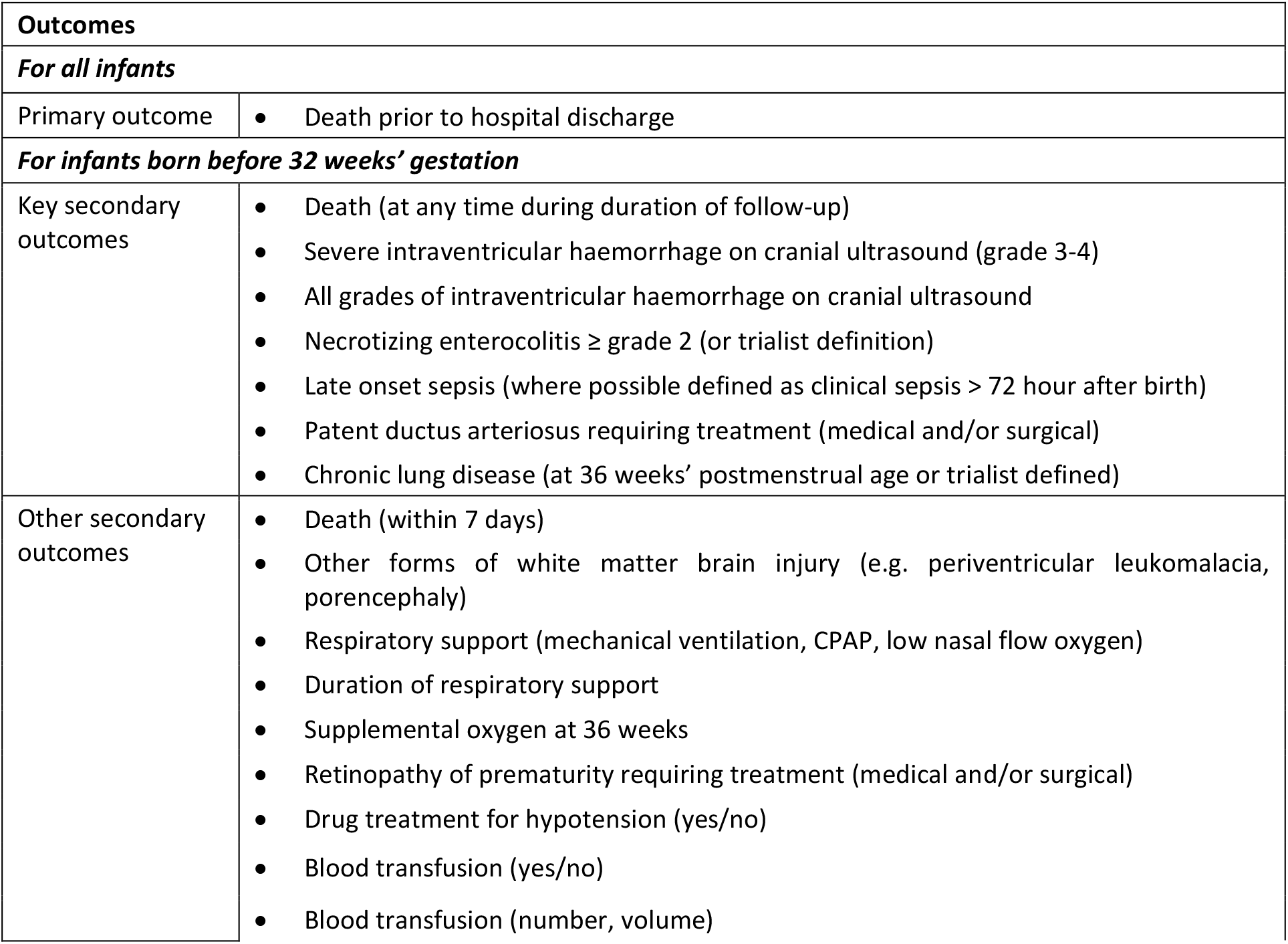
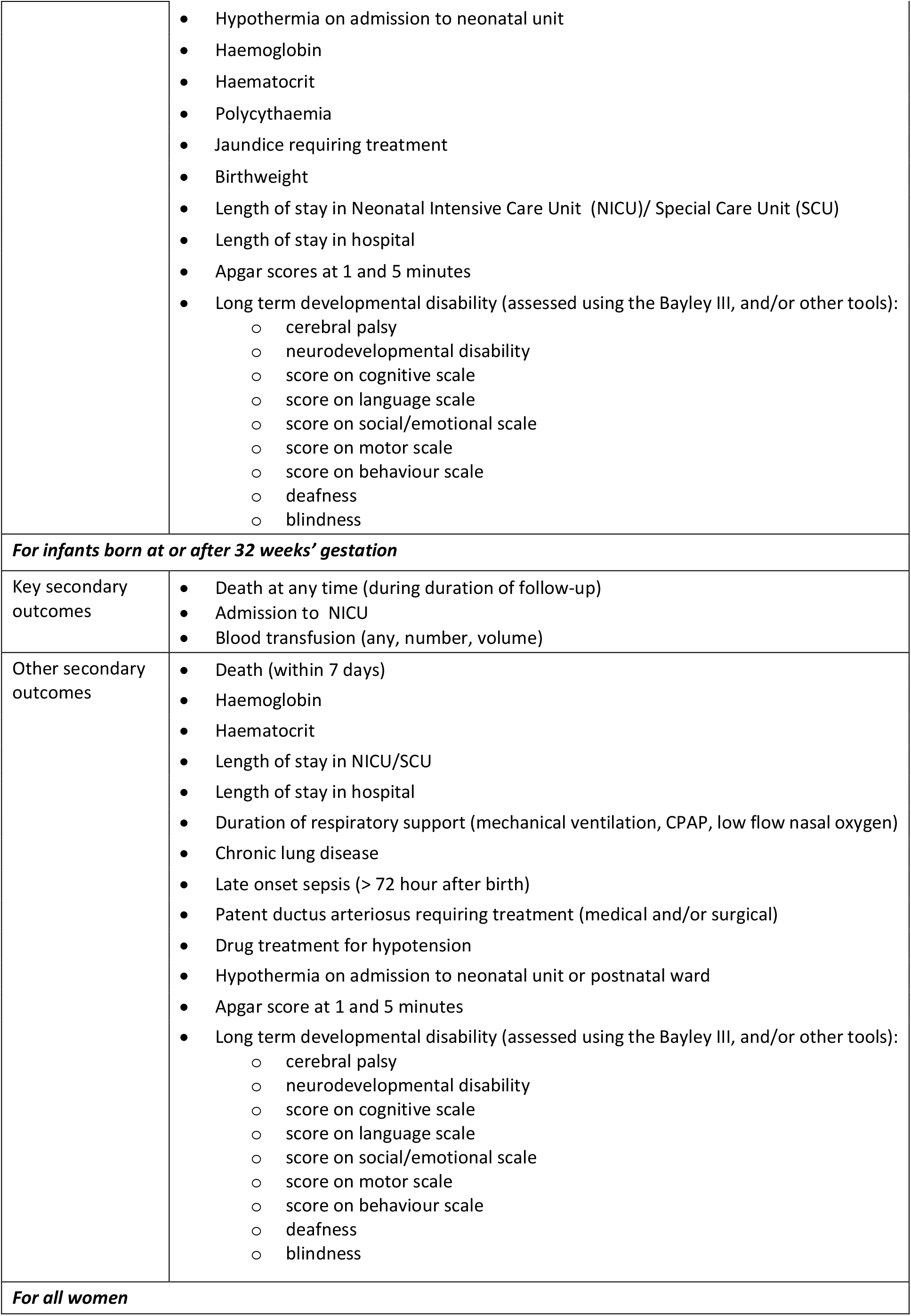
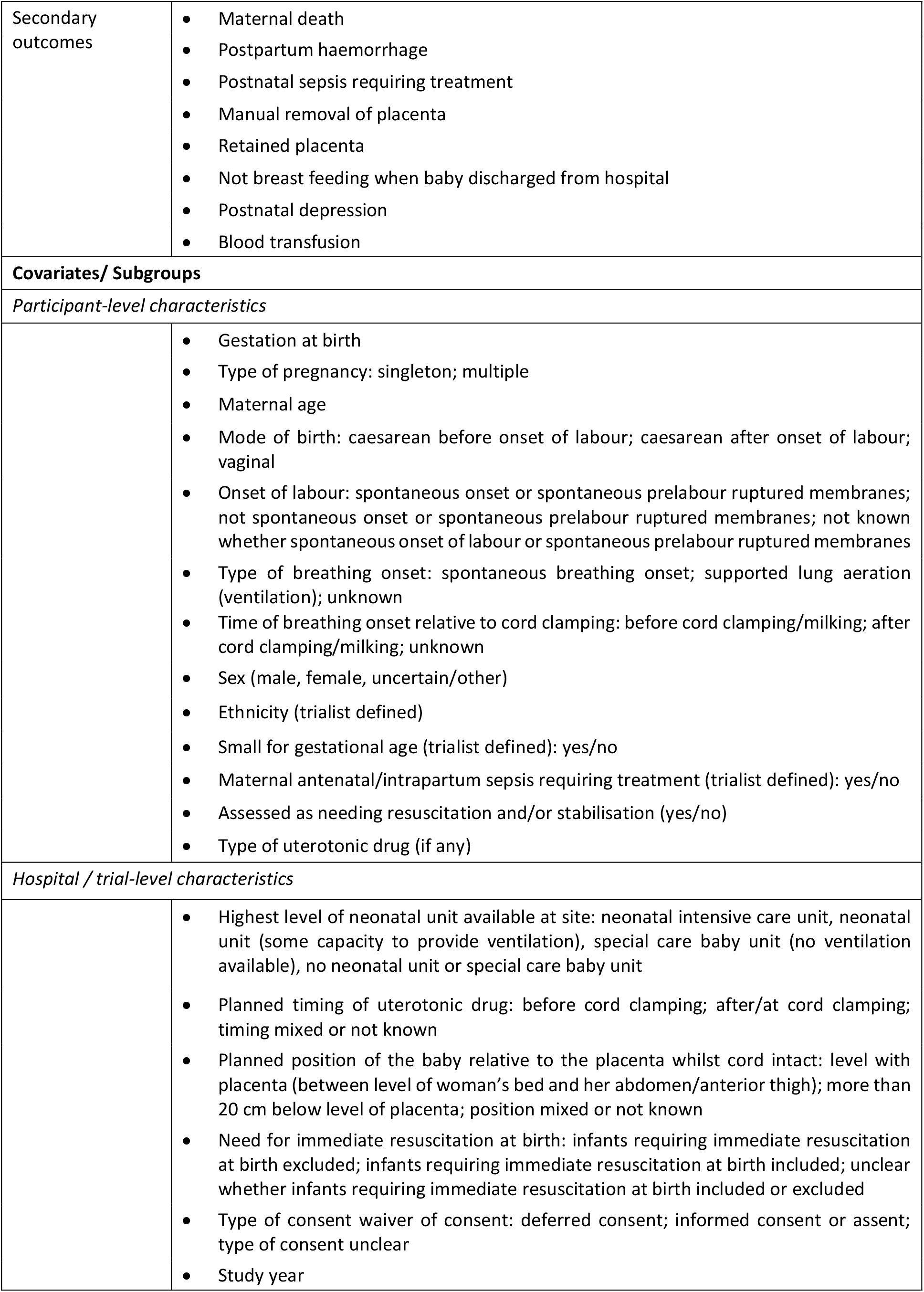
Measures for individual participant data pairwise meta-analysis.

### Covariates and subgroups for pairwise meta-analysis

Subgroup analyses will be conducted for the primary outcome of death (prior to hospital discharge) and two key secondary outcomes (IVH any grade and any infant blood transfusion) if sufficient data are available. All included covariates and subgroups are listed in Table 1. The comparative effects of alternative cord management strategies may vary depending on key infant risk factors, and/or on the level and type of neonatal care available at the hospital of birth. Thus, there will be subgroup analyses based on participant level characteristics and based on hospital level characteristics. If data are insufficient for the pre-specified subgroup analyses, categories will be collapsed.

### Data analysis for pairwise meta-analysis

The full Statistical Analysis Plan will be agreed on by the iCOMP Collaboration before any analyses are undertaken. Analyses will include all randomised participants for which data are available, and the primary analyses will be based on intention-to-treat. Analyses will be conducted using the open-source software R.(56)

For each outcome, a one-stage approach to analysis will be employed to include individual participant data from all eligible trials in a multilevel random or mixed effects regression model. Aggregate data will be included were individual participant data are unavailable.(57) Relative heterogeneity of treatment effects across trials will be estimated using I^2^, with further inclusion in secondary models of participant level and trial level covariates to explain the sources of heterogeneity. Prediction intervals will be estimated to ascertain absolute heterogeneity. Forest plots will be presented by trial for the primary outcome, and for any secondary outcomes where there is evidence of heterogeneity across trials.

We will use a generalised linear modelling framework, with the choice of outcome distribution and link function dependent on outcome type. For example, binomial with log link will be used to estimate risk ratios for binary outcomes, and Gaussian with identity link for mean differences, with log-transformation of the data if appropriate. We will follow a similar approach for secondary outcomes. For estimation of subgroup effects, we will present forest plots of pooled treatment effects according to pre-specified subgroup variables, and estimate effects by including appropriate interaction terms between a subgroup variable and treatment arm in the regression models. The results of all comparative analyses will be presented using appropriate estimates of treatment effect along with 95% confidence intervals and two-sided p-values.

In advance of conducting the analyses, we will decide whether there are sufficient reliable data to allow meaningful analysis of any individual outcome or subgroup. If not, the analysis will not be conducted, and this will be reported subsequently.

### Outcome measures for network meta-analysis

The primary outcome for the network meta-analysis will be death of the baby during the initial hospital stay. If data availability permits, IVH (any grade) and blood transfusion (any) will be analysed as two key secondary outcomes.

### Covariates and subgroups for network meta-analysis

Gestational week at birth and highest level of available care will be considered as effect modifiers to improve consistency of the NMA model. There will be subgroup analyses assessing treatment effect by week of gestational age, and by comparing babies assessed as in need of immediate resuscitation versus not in need of immediate resuscitation.

### Assessment of the transitivity assumption for network meta-analysis

Transitivity in the network will be assured by only including interventions that are regarded as jointly randomisable and by limiting our sample to preterm infants. Gestational age at birth, hospital setting (highest level of available neonatal care), as well as study year may act as effect modifiers and could influence the transitivity of the network. We will therefore investigate whether these variables are distributed evenly across comparisons. If we find any of those variables to be unevenly distributed, they will be included in the network as covariates to investigate their influence on the network and on possible inconsistency.

### Data analysis for network meta-analysis

As for the pairwise meta-analysis, all analyses will be specified a-priori in a full Statistical Analysis Plan, all randomised participants for which data are available will be included, and the primary analyses will be intention-to-treat.

We will calculate a two-step random-effects contrast-based network meta-regression to compare and rank all available interventions for the primary outcome death (during initial hospital stay) and, if data permits, for the two key secondary outcomes - IVH (any grade) and blood transfusion (any). Summary risk ratios with confidence and prediction intervals will be presented for each pairwise comparison in a league table. We will estimate the ranking probability of each intervention being at each rank, and we will use surface under the cumulative ranking curve (SUCRA) and mean ranks to obtain a treatment hierarchy. A frequentist approach to analysis will be used. Should models not converge, a Bayesian approach will be used instead, setting a prior of no effect and a large variance. Correlations induced by multi-arm studies will be accounted for using multivariate distributions.

As a second step, interactions between key covariates and effect estimates will be tested, assuming a common interaction across comparisons. If there are statistically significant interactions between covariates and treatment effects, we will provide probability rankings of intervention effects by subgroup for these covariates. A single heterogeneity parameter will be assumed for each network.

### Assessment of inconsistency for network meta-analysis

Global consistency will be assessed using the Q statistics for inconsistency and the design-by-treatment interaction model. Local consistency will be assessed using the loop-specific approach and the node-splitting approach to explore sources of inconsistency. Since tests of inconsistency are known to have low power,(58) results will be interpreted with caution, and potential known sources for inconsistency will be explored even if there is no statistical evidence of inconsistency. Any detected inconsistency will be explored by including covariates into the model, and by excluding potential outlier studies in sensitivity analyses. A judgement of excessive heterogeneity or inconsistency would prevent the interpretation and reporting of the network meta-analysis.

### Assessment of compliance with the allocated intervention

Compliance with the interventions will be described for each trial. For studies of early versus deferred cord clamping this will be based on i) the time to cord clamping in each allocated group, and ii) the difference in time between early and deferred clamping. For studies comparing cord milking with no milking, this will be based on i) time to cord clamping in the allocated groups, and ii) reported compliance with cord milking in both groups.

### Assessment of selection bias

We will perform a nested prospective meta-analysis as a sensitivity analysis, to detect potential differences between prospectively and retrospectively included studies that may point to selection or publication bias. We expect to also be able to include some unreported outcomes sourced from the individual participant data provided by the included studies, alleviating selective outcome reporting bias. Additionally, comparison-adjusted and contour-enhanced funnel plots(59) will be utilised to examine whether there are differences in results between more and less precise studies.

### Adjustments for multiple testing

There is only one primary outcome, and few key secondary outcomes for this study. For other secondary outcomes, no formal adjustments for multiple testing are planned but instead, we will be following the approach outlined by Schulz and Grimes(60): as secondary outcomes examined in this study are interrelated, we will interpret the pattern of results, examining consistency of results across related outcomes, instead of focusing on any single, statistically significant result. All secondary outcomes will be reported. Subgroup analyses will be performed by testing for interactions and findings will be reported as exploratory.(61)

### Planned sensitivity analyses

To assess whether results are robust to trial characteristics and methods of analysis, the following sensitivity analyses will be conducted for the primary outcome, if data are sufficient:

- Excluding studies with high risk of bias for sequence generation and/or concealment of allocation and/or loss to follow up for pairwise and network meta-analysis;
- For trials comparing early cord clamping with deferred clamping, analysis of outcomes weighted by degree of separation in mean actual timing of cord clamping between intervention and control groups for pairwise meta-analysis;
- Analysis of outcomes weighted by degree of separation in haemoglobin (at 24 hours) achieved between intervention and control groups for pairwise meta-analysis (as a surrogate for net placental transfusion);
- For trials with deferred cord clamping, an additional dose-response analysis assessing intended time of cord clamping deferral as a continuous variable will be performed;
- Exploratory analysis based on actual, rather than intended, timing of cord clamping for individual participants for pairwise and network meta-analysis;
- The impact of missing data on the effects of the included interventions for the primary outcome may be explored (if appropriate).

### Project management

The iCOMP Collaboration will invite membership from representatives of each of the included trials contributing individual participant data, have a Secretariat, and invite methodological and clinical experts who will form an Advisory Group. The Secretariat will be responsible for data collection, management and analysis, and for communication within the Collaboration.

### Ethical issues

For each included trial, ethics approval has been previously granted by their respective Human Research Ethics Committees (or equivalent), and informed consent has been obtained from all participants. The Chief Investigators of the included trials remain the custodians of their own trial’s data. Individual participant data from the included trials will be de-identified before sharing with the iCOMP Collaboration. Ethics approval for this project has been granted by the University of Sydney Human Research Ethics Committee (number: 2018/886).

### Publication policy

The key methods for this meta-analysis protocol were agreed by the iCOMP Collaborators in January 2015, before unblinding of any outcome data from the studies included in the nested prospective meta-analysis. This manuscript was discussed at the iCOMP Collaborators’ meeting held at the Pediatric Academic Societies meeting in San Diego in April 2015. At this meeting it was agreed the protocol should be expanded to include a retrospective systematic review and individual participant data and network meta-analysis with a nested prospective meta-analysis. The protocol was then revised, based on further discussion, and circulated to members of the iCOMP Collaboration for further comment and agreement prior to manuscript submission.

Participating trialists in the prospective meta-analysis, when reporting results from their own trials, will endeavour to include a statement that their trial is part of this prospective meta-analysis in any published manuscripts or conference abstracts. Any reports of the results of this meta-analysis will be published either in the name of the collaborative group, or by representatives of the collaborative group on behalf of the iCOMP Collaboration, as agreed by members of the collaborative group. Draft reports will be circulated to the collaborative group for comment and approval before submission for publication.

## DISCUSSION

There is an urgency to conduct this systematic review and pairwise individual participant data and network meta-analysis so we can make sense of the numerous trials currently being undertaken, inform clinical practice, and identify the most promising interventions for further evaluation.

This meta-analysis offers an opportunity to reliably test important hypotheses that cannot be resolved by any of the individual trials, either alone or in simple combination. Coordinating international efforts in this way will help achieve consensus on the most important substantive clinical outcomes to assess in any future trials as needed. Unequivocal synthesised results, together with the identification of key determinants (e.g. effect modifiers), will be critical for translating evidence from this meta-analysis directly into practice. Figure 2 shows the network of comparisons available from the trials identified to date. We plan to complete study identification and individual participant data collection by end-2019, then conduct the analyses and disseminate results by mid-2021.

**Figure 2.**
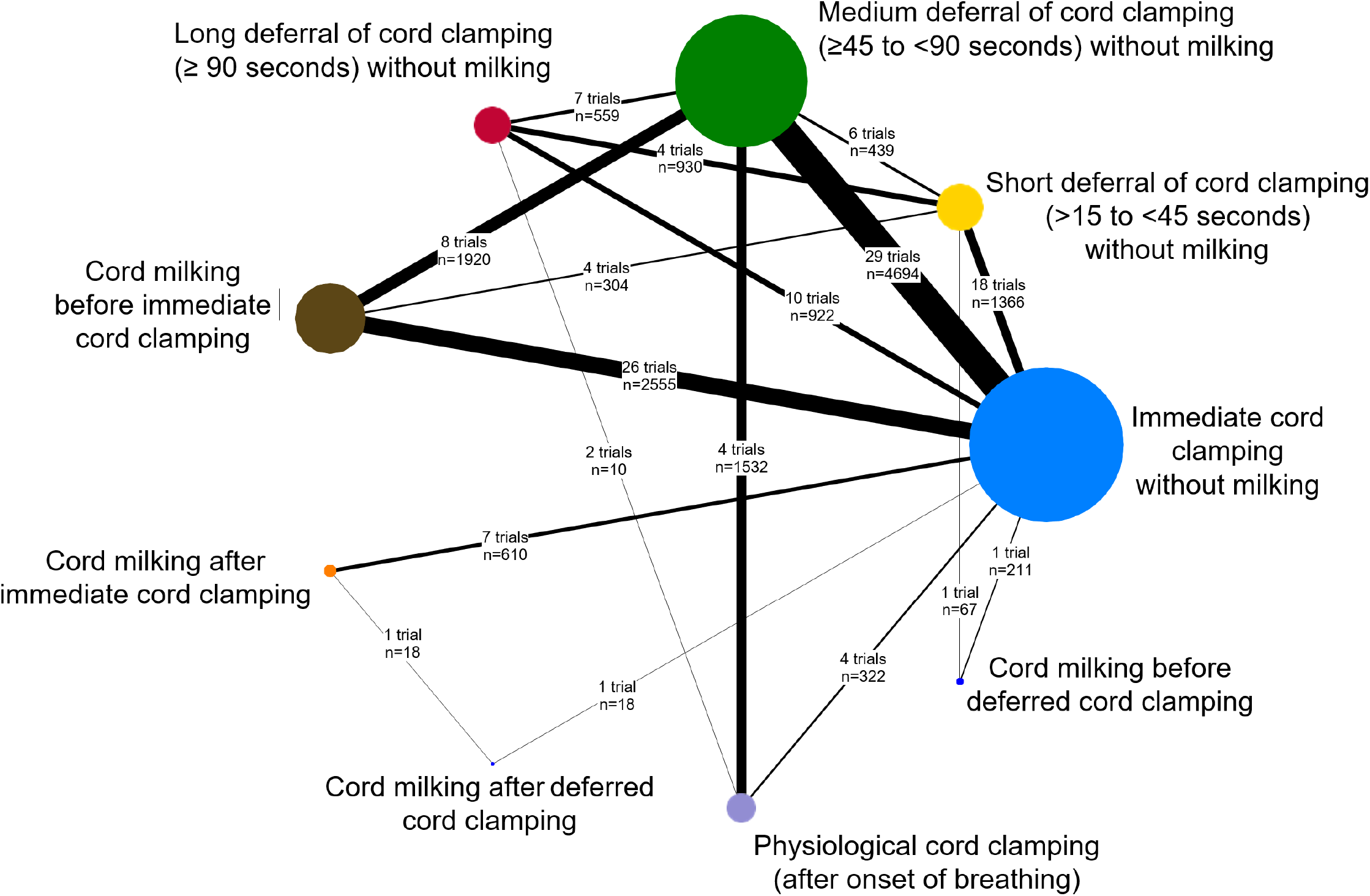
Illustration of network of currently available trials comparing different timing of cord clamping.

This study is only possible because trialists around the world have agreed to collaborate to share the individual participant data from their cord management trials. This collaborative approach will enable us to move beyond the traditional “one-size-fits-all” and towards precision medicine, to find the optimal intervention from a range of treatment options for each individual woman and her baby, based on their individual characteristics and risk factors.

## Data Availability

This is a protocol and therefore no data are currently available.

## Acknowledgements

Thanks to Sarah Somerset, Min Yang, Charlotte Lloyd, and Virginia Portillo for previous support for the Secretariat and input into earlier protocol drafts.

## Authors’ contributions

LD and LA conceived the idea. ALS, LA, and LD drafted the protocol with input from all authors. All authors have agreed the final manuscript. ALS is the guarantor of the review.

## List of abbreviations

iCOMP: **i**ndividual participant data on **Co**rd **M**anagement at **P**reterm birth
CENTRAL: Cochrane Central Register of Controlled Trials
CINeMA: Confidence in Network meta-analysis
CPAP: continuous positive airway pressure
IPD: individual participant data
NICE: National Institute for Health and Care Excellence
NICU: neonatal intensive care unit
NIHR: National Institute for Health Research
NMA: network meta-analysis
PRISMA: Preferred Reporting Items for Systematic Review and Meta-Analysis
UK: United Kingdom
WHO: World Health Organization
ILCOR: International Liaison Committee on Resuscitation
PROSPERO: International prospective register of systematic reviews

**iCOMP collaboration members:**

**<u>Secretariat:</u>** Anna Lene Seidler^1^, Lelia Duley^2^, Alan Montgomery^2^, Kylie E Hunter^1^, Angie Barba^1^, Ava Grace Tan-Koay^1^, Lisa Askie^1^

**<u>Trialists:</u>** Amir Kugelman^3^, Anu George^4^, Anu Sachdeva^5^, Anup Katheria^6^, Arjan Te Pas^7^, Ashish Kc^8^, Bimlesh Kumar^9^, Carl Backes^10^, Catalina De Paco Matallana^11^, Chamnan Tanprasertkul^12^, Chayatat Ruangkit^13^, Eugene Dempsey^14^, G Ram Mohan^15^, Guillermo Carroli^16^, Heidi Al-Wassia^17^, Hytham Atia^18^, Heike Rabe^19^, Islam Nour^20^, Jiangqin Liu^21^, John Bauer^22^, John Kattwinkel^23^, Judith Mercer^24^, Justin Josephsen^25^, Karen Fairchild^23^, Kellie Murphy^26^, Kristy Robledo^1^, Lakhbir Dhaliwal^15^, Laura Perretta^27^, Lin Ling^28^, Manoj Varanattu^29^, Maria Goya^30^, Musa Silahli^31^, Neelam Kler^32^, Neil Finer^33^, Ola Andersson^8^, Omar Kamlin^34^, Peter Giannone^22^, Pharuhad Pongmee^35^, Prisana Panichkul^36^, Sandeep Kadam^37^, Sangkae Chamnanvanakij^36^, Shigeharu Hosono^38^, Shiraz Badurdeen^34^, Thomas Ranjit^39^, Venkataseshan Sundaram^15^, Victor Lago Leal^40^, Vikram Datta^41^, Waldemar Carlo^42^, Walid El-Naggar^43^, William Tarnow-Mordi^1^, William Oh^44^

**<u>Advisers:</u>** Graeme Polglase^45^, John Simes^1^, Martin Kluckow^46^, Peter Davis^34^, Stuart Hooper^45^, Thomas P.A. Debray^47^.

^1^ National Health and Medical Research Council Clinical Trials Centre, University of Sydney, Sydney, Australia

^2^ Nottingham Clinical Trials Unit, University of Nottingham, Nottingham, UK

^3^ Bnai Zion Medical Center, Haifa, Israel

^4^ Malankara Orthodox Syrian Church Medical College, Kerala, India

^5^ All India Institute of Medical Science, New Delhi, India

^6^ Sharp Healthcare, San Diego, California, USA

^7^ Leiden University, Leiden, Netherlands

^8^ Uppsala University, Uppsala, Sweden

^9^ Lala Lajpat Rai Memorial Medical College, Uttar Pradash, India

^10^ Ohio State University Wexner Medical Center, Columbus, Ohio, USA

^11^ Clinic University Hospital Virgen de la Arrixaca, Murcia, Spain

^12^ Thammasat University, Pathumthani, Thailand

^13^ Mahidol University, Samut Prakan, Thailand

^14^ Cork University Maternity Hospital, Cork, Ireland

^15^ Post Graduate Institute of Medical Education & Research, Chandigarh, India

^16^ Rosarino Center for Perinatal Studies, Rosario, Argentina

^17^ King Abdulaziz University, Jeddah, Saudi Arabia

^18^ Zagazig University, Zagazig, Egypt

^19^ Brighton and Sussex Medical School, University of Sussex, Brighton, UK

^20^ Mansoura University Children’s Hospital, Mansoura, Egypt

^21^ Tongji University School of Medicine, Shanghai, China

^22^ University of Kentucky, Lexington, Kentucky, USA

^23^ University of Virginia, Charlottesville, Virginia, USA

^24^ The University of Rhode Island, Kingston, Rhode Island, USA

^25^ St Louis University School of Medicine, St. Louis, Missouri, USA

^26^ Mount Sinai Hospital, Toronto, Canada

^27^ Weill Cornell Medical College, New York City, New York, USA

^28^ Suining Central Hospital, Sichuan, China

^29^ Jubilee Mission Medical College & Research Centre, Kerala, India

^30^ Vall d’Hebron University Hospital, Barcelona, Spain

^31^ Baskent University, Ankara, Turkey

^32^ Sir Ganga Ram Hospital, New Delhi, India

^33^ University of California, San Diego, California, USA

^34^ The Royal Women’s Hospital, Melbourne, Australia

^35^ Ramathibodi Hospital, Mahidol University, Bangkok, Thailand

^36^ Phramongkutklao Hospital, Ratchathewi, Bangkok, Thailand

^37^ KEM Hospital, Pune, Maharashtra, India

^38^ Jichi Medical University Saitama Medical Center, Saitama, Japan

^39^ St John’s Medical College & Hospital, Bangalore, Kamataka, India

^40^ University Hospital of Getafe & European University of Madrid, Madrid, Spain

^41^ Lady Hardinge Medical College, New Delhi, India

^42^ University of Alabama at Birmingham, Birmingham, Alabama, USA

^43^ Department of Pediatrics, Dalhousie University, Halifax, Canada

^44^ Women and Infants’ Hospital, Providence, Rhode Island, USA

^45^ The Ritchie Centre, Obstetrics & Gynaecology, Monash University, Melbourne, Australia

^46^ Faculty of Medicine and Health, University of Sydney, Sydney, Australia

^47^ Julius Center for Health Sciences and Primary Care, University Medical Center Utrecht, Utrecht, the Netherlands

